# Dopaminergic medication increases motivation to exert cognitive control by reducing subjective effort costs in Parkinson’s patients

**DOI:** 10.1101/2022.02.07.22270623

**Authors:** Mario Bogdanov, Sophia LoParco, A. Ross Otto, Madeleine Sharp

## Abstract

Engaging in demanding mental activities requires the allocation of cognitive control, which can be effortful and aversive. Individuals thus tend to avoid exerting cognitive effort if less demanding behavioral options are available. Recent accounts propose a key role for dopamine in motivating behavior by increasing the sensitivity to rewards associated with effort exertion. Whether dopamine additionally plays a specific role in modulating the sensitivity to the costs of cognitive effort, even in the absence of any incentives, is much less clear. To address this question, we assessed cognitive effort avoidance in patients (n = 38) with Parkinson’s disease, a condition characterized by loss of midbrain dopaminergic neurons, both ON and OFF dopaminergic medication and compared them to healthy controls (n = 24). Effort avoidance was assessed using the Demand Selection Task (DST), in which participants could freely choose between performing a high-demand or a low-demand version of a task-switching paradigm. Critically, participants were not offered any incentives to choose the more effortful option, nor for good performance. Healthy controls and patients OFF their dopaminergic medications preferred the low-demand option, in keeping with the tendency to avoid effort on this task previously demonstrated in young adults. In contrast, patients ON dopaminergic medications displayed significantly less effort avoidance than when they were OFF medications. This change in preference could not be explained by differences in task-switching performance or the patients’ ability to detect the different levels of cognitive demand in the DST. Our findings provide evidence that dopamine replacement in Parkinson’s patients increases the willingness to engage in cognitively demanding behavior, even in the absence of any clear benefits. These results suggest that dopamine plays a role in reducing the sensitivity to effort costs that is independent of its role in enhancing the sensitivity to the benefits of effort exertion.

## 1. Introduction

Adaptive, goal-directed behavior requires the engagement of cognitive control, for example when attenuating distracting noise during remote work or when keeping track of multiple ongoing projects and deadlines. Given the limitations of our cognitive capacity, however, employment of cognitive control is costly and subjectively effortful, necessitating a strategic allocation of cognitive resources to goals that are worth the investment (Inzlicht et al., 2018; Kool & Botvinick, 2018; Kurzban et al., 2013). On this view, recent research has demonstrated that an individual’s decision about whether to engage in a cognitively demanding behavior is governed by a cost-benefit trade-off that weighs the anticipated degree of effort against the subjective value of the prospective reward (Kurzban et al., 2013; Otto & Daw, 2019; Shenhav et al., 2017; Westbrook & Braver, 2015). Akin to the well-known “law of least work” postulated by Hull (1943), people will avoid cognitively effortful behavior if a comparable reward can be obtained by putting in less mental work (Bogdanov et al., 2021; Kool et al., 2010; Patzelt et al., 2019). Echoing converging findings in animal and human research on physical effort (Denk et al., 2005; Floresco et al., 2008; Mazzoni et al., 2007; Pasquereau & Turner, 2013; Salamone et al., 2009, 2016; Tanaka et al., 2021; Treadway, Buckholtz, et al., 2012; Varazzani et al., 2015), mechanistic accounts of cognitive control emphasize the role of striatal and prefrontal dopamine not only in supporting higher-order cognitive processes, but also in motivating cognitive effort exertion by modulating the cost-benefit computations presumably directing effort allocation decisions (Cocker et al., 2012; Cools, 2016; Westbrook et al., 2020; Westbrook & Braver, 2016). In line with this idea, pharmacological manipulations to increase synaptic dopamine concentrations have been recently demonstrated to bias human subjects towards increased willingness to exert high levels of cognitive effort for larger rewards, and this has been shown across several cognitive processes and task domains, including attention, working memory, and task-switching (Hofmans et al., 2020; Manohar et al., 2015; Timmer et al., 2018; Westbrook et al., 2020).

Taken together, these findings suggest that the dopaminergic system is critically important in overcoming the subjective aversion to cognitive effort, but it is less clear if this depends on the concomitant evaluation of the costs and benefits conferred by effort exertion. Indeed, the bulk of empirical work on the role of dopamine in effort-based decision-making has relied on experimental paradigms that explicitly manipulate both the degree of effort required to obtain a reward as well as the magnitude of the reward itself, making it difficult to assess the specificity of these effects. While there is some evidence that dopamine may be primarily involved in signaling upcoming rewards instead of effort costs (Walton & Bouret, 2019; Westbrook et al., 2020), a recent study in young, healthy adults demonstrated that administration of methylphenidate, which increases catecholamine levels in the brain, may decrease the subjective aversiveness of cognitive effort even in the absence of additional incentives, although the strength of the effect was dependent on participants’ trait impulsivity (Froböse et al., 2018). However, given that methylphenidate affects both dopamine and noradrenaline, it remains unclear whether dopamine in particular attunes motivational processes primarily by increasing sensitivity to the rewarding outcomes of effort exertion instead of, or in addition to, reducing sensitivity to the costs of effort.

This question is especially pertinent to Parkinson’s disease, a neurodegenerative disorder characterized by the loss of dopaminergic neurons in the substantia nigra (pars compacta). A large proportion of patients affected by Parkinson’s Disease suffer from apathy and/or anhedonia, extreme forms of amotivation that drastically affect patients’ quality of life and disease prognosis (den Brok et al., 2015; Husain & Roiser, 2018; Lemke et al., 2005; Treadway, Bossaller, et al., 2012). Only a few studies have sought to systematically examine the factors that influence effort exertion in Parkinson’s disease. In keeping with results from pharmacological studies in healthy adults, initial reports demonstrate increased motivation to exert both physical and cognitive effort in exchange for larger reward when patients were ON compared to OFF their dopaminergic medication (Chong et al., 2015; McGuigan et al., 2019). However, given the evidence for dopamine-dependent reduced reward sensitivity in Parkinson’s disease (Aarts et al., 2012; Bódi et al., 2009; Brown et al., 2020; Pilgrim et al., 2021; Schott et al., 2007; Sharp et al., 2015, 2020), it is especially important to consider the influence of dopaminergic medications on effort in isolation of its effects on reward.

As of yet, no study has investigated whether dopaminergic medications affect individuals’ fundamental willingness to engage in cognitively effortful behavior in the absence of additional incentives in Parkinson’s disease. Here, we aimed to address this question by examining the role of dopamine in modulating the tendency towards effort avoidance in a well-established effort-preference task, termed the demand selection task (DST; Gold et al., 2015; Kool et al., 2010). In the DST, participants repeatedly choose between two options: one that most often leads to a low cognitive demand task and one that leads to a high cognitive demand task, where level of cognitive demand is determined by the frequency of task switches in a task switching paradigm (da Silva Castanheira et al., 2021; Dreisbach & Haider, 2006; Liu & Yeung, 2020; Monsell, 2003). Critically, unlike in the tasks used in past studies on effort-based decision-making in Parkinson’s disease (Chong et al., 2015; McGuigan et al., 2019), there were no incentives on offer which would influence participants’ choice between the low-versus high-demand options. Thus, participants’ choices in this task should solely reflect their sensitivity to the costs of cognitive effort independent of their sensitivity to rewards. Indeed, previous work using the DST shows that, on average, participants prefer the low-demand cognitive task—or, alternatively, avoid the high-demand task (Bogdanov et al., 2021; Froböse et al., 2018; Patzelt et al., 2019). To investigate how dopamine affects effort avoidance, we tested Parkinson’s patients both ON and OFF their usual medication as well as age-matched healthy controls in a two-day mixed design. Based on earlier findings (Froböse et al., 2018; McGuigan et al., 2019), we hypothesized patients OFF dopamine would exhibit more effort-avoidant preferences, compared to when they were ON medication, as well as compared to healthy controls. In addition, we expected that patients ON medication would show similar rates of effort avoidance than healthy controls. Finally, the task-switching paradigm embedded in the DST allowed us to explore a possible relationship between participants’ switch costs—as a proxy for individual effort costs—and effort-avoidant choice behavior in the DST.

## 2. Material and methods

### 2.1 Participants

A total of 68 participants were recruited for the study: 42 Parkinson’s patients (14 females, mean ± SD age: 64.24 ± 6.76 years) and 26 healthy, age-matched controls (22 females, mean ± SD age: 62.64 ± 7.97). Patients were recruited from the Movement Disorder Clinic at the Montreal Neurological Institute, community groups, and from the Quebec Parkinson Network, a registry of patients with Parkinson’s disease interested in research who have been referred by movement disorder specialists. Control participants were recruited from spouses and friends of patients, community groups, and social media posts. None reported major health issues, neurological disorders, or active psychiatric problems. Disease duration in patients ranged from 0.42 to 14.5 years (mean ± SD age: 5.65 ± 4.18). All patients were taking levodopa. All subjects gave informed written consent and were compensated for their participation. The study was approved by the McGill University Health Centre Research Ethics Board and all procedures were performed in accordance with the appropriate institutional guidelines.

After examining response patterns in the DST, we found that six participants (4 Parkinson’s patients and 2 healthy controls) evidenced difficulty understanding the task instructions, demonstrated by 0% correct responses in the final two instructed blocks of the DST (see below). These participants were excluded from the analysis, leaving a sample of n =62 (38 patients and 24 healthy controls). Demographic information of this final sample is depicted in Table 1.

**Table 1.** Sample demographics and neuropsychological assessment. ***Note***. LEED = Total Levodopa equivalent dose, MoCA = Montreal Cognitive Assessment, verbal fluency is taken from the language section of the MoCA, SDMT = Symbol digit modalities test, GDS = Geriatric Depression Scale, AES = Apathy Evaluation Scale. Values are presented as mean ± SD. \**p* < .05, \*\**p* < .01, *** *p* < .001.

|  | Parkinson's patients | Healthy controls | *p*-value |
| --- | --- | --- | --- |
| Age | 64.74 ± 7.35 | 63.59 ± 6.92 | .564 |
| Education (years) | 15.47 ± 3.83 | 15.95 ± 2.36 | .578 |
| Disease duration (years) | 5.43 ± 3.95 | / | / |
| LEED (mg) | 649.38 ± 333.84 | / | / |
| Percent female | 37% | 92% | <.001*** |
| MoCA | 27.77 ± 1.41 | 28.35 ± 1.37 | .139 |
| Verbal fluency (MoCA) | 13.65 ± 3.92 | 15.23 ± 3.26 | .116 |
| Digit span forward | 6.48 ± 1.43 | 6.77 ± 1.41 | .468 |
| Digit span backward | 5.13 ± 1.23 | 5.14 ± 1.39 | .971 |
| SDMT | 40.84 ± 10.97 | 48.41 ± 6.72 | .003** |
| GDS | 8.37 ± 6.27 | 5.32 ± 4.92 | .055 |
| AES | 58.87 ± 7.74 | 60.62 ± 7.17 | .408 |

### 2.2 Procedure and design

All participants were tested in a two-session, within-subject design. The interval between both testing days was at least 6 weeks in order to minimize practise effects. Testing sessions started in the morning between 9 and 12 a.m. to control for the timing of medication intake and circadian factors. Parkinson’s patients were either tested one hour after having taken their dopaminergic medication (ON session) or after an overnight withdrawal (minimum 15h) from their medication (OFF session). The order of ON and OFF sessions was counterbalanced across Parkinson’s patients. Ten patients withdrew from the experiment after the first day of testing: eight patients missed their OFF session; two patients missed their ON session. Reasons were either severe motor symptoms during the OFF state (n = 2) or not otherwise specified (n = 8). Similarly, nine healthy controls missed their second session. This was due to the start of the Covid-19 pandemic that terminated data collection early. Finally, due to technical reasons, two healthy controls only have valid data for their second testing session. All of these subjects with a single session were still included in our final analyses.

Participants were also administered a neuropsychological battery on the first session to establish baseline cognitive functioning. The neuropsychological battery consisted of the Montreal Cognitive Assessment (MoCA), the Digit Span (forward and backward), and the Symbols Digit Modalities Test (SDMT). In addition, participants completed the Geriatric Depression Scale (GDS) and the Apathy Evaluation Scale (AES). Results from Welch’s two-sample t-tests indicate that Parkinson’s patients performed similarly to healthy controls in most measures but scored significantly lower in the SDMT (*p* = .003; see Table 1).

### 2.3 Task-switching paradigm

On both testing sessions, participants completed 125 trials of a simple task switching paradigm (Gold et al., 2015; Kool et al., 2010). On each trial, participants were presented with a colored number between 1 and 9 (excluding 5). Based on its color (blue or yellow), participants had to indicate by button press whether the number they saw was either larger or smaller than 5 (magnitude judgment) or whether it was odd or even (parity judgment). The mapping between colors and button presses was counterbalanced between participants. Critically, the color of the number (and therefore the judgment task participants had to perform), switched from trial to trial with a probability of 50%. We reasoned that participants with greater switch costs (i.e., greater reduction in accuracy and slowing on trials that represent a switch) may experience the task as more effortful, which may lead them to be more effort averse in the DST (Kool et al., 2010). By establishing baseline switch costs, we thus aimed to control for such individual (and potential disease or medication) differences that might affect choice behavior in the DST (see below).

### 2.4 Demand Selection Task (DST)

The demand selection task (DST) was used to measure participants’ level of effort avoidance by making them choose between engaging in two distinct difficulty levels of a task switching paradigm similar to the one described above. The protocol used in the present study was adapted from a procedure described in previous work (Bogdanov et al., 2021; Gold et al., 2015; Experiment 3 in Kool et al., 2010). On each trial of this task, participants were instructed to choose between two abstract multicolor patches presented on screen. To make their selection participants moved the mouse cursor over their preferred patch, which revealed either a blue or a yellow number between 1 and 9 (excluding 5). As above, the color of the number indicated the type of judgment (magnitude or parity) participants had to make. Critically, the two patches differed in their task-switching rate, i.e., the frequency in which their associated numbers would change color (and therefore judgement type) compared to the previous trial. More specifically, for the low cognitive demand patch, the color of the number would repeat with a probability of 90% whereas for the high-demand patch, the probability of color repetition was only 10%, requiring more task-switching and thus more cognitive effort. Importantly, participants were not explicitly informed about this difference. Instead, participants were instructed to freely choose between both color patches, that they might notice differences between them and that, if they developed a preference for one over the other, they could choose their preferred patch more frequently. It should be noted here that participants were not incentivized for better performance or for choosing the harder task. As such, deviations from purely random choice behavior in the DST should reflect an individual’s preference for high or low task demand.

Participants performed four of these free choice blocks (50 trials/block) for a total of 200 trials. A new pair of color patches presented in a new location on screen was used for each block. Following Gold et al., 2015, we included two additional forced choice blocks in our DST. On these blocks, participants were informed explicitly that one patch led to a more difficult task than the other because of more frequent switches and they were specifically instructed to choose either the easier (block 5) or more difficult (block 6) patch. This was done to measure whether participants were able to detect the differences cognitive effort demands and to control for the possibility that group differences in free choice behavior could be caused by differences in detection ability rather than effort aversion. The two forced choice blocks consisted of 35 trials each. The DST was programmed using PsychToolbox (Kleiner et al., 2007) for MATLAB (The MathWorks, Natick, MA).

### 2.5 Statistical Analysis

All analyses were performed using the lme4 package (Version 0.999375-32; Bates et al., 2014) in the R programming environment (Version 3.2.6; R Core Team, 2020), with the critical alpha level set at *p* = .05. We used linear and logistic mixed effects regression models to analyse our main dependent variables of interest. In order to compare groups we defined two binary, dummy-coded variables to represent the between-subject effect of disease state (0= Parkinson’s patients, 1 = healthy controls) and the within-subject effect of dopaminergic drug state (0 = OFF dopaminergic medication, 1 = ON dopaminergic medication). In this operationalization, patients OFF dopamine are the designated baseline group such that the effect of disease state reflects group differences between controls and patients OFF and the drug state effect reflects the difference between patients ON and OFF (Gelman & Hill, 2006; Sharp et al., 2015).

To analyze choice behavior in the DST, we performed a mixed effects logistic regression that predicted low-demand choices (0 = high demand option chosen, 1 = low demand option chosen) in the free choice blocks as a function of disease state, drug state, trial number within a given block, and session (effect coded: -1 = first session, 1 = second session). Trial number was included to examine the time course of effort avoidant choice behavior over time, as participants were presented with new color patches every block.

Session was included to control for potential training effects across testing days. We also included interaction terms for disease state and session, disease state and trial number, drug state and session, as well as drug state and trial number in the model. Participants’ choice reaction times (i.e., the time it took them to choose between the color patches) were analyzed in a linear mixed effects regression using the same predictors. To analyze task-switching performance during the DST (i.e., accuracy and correct RTs) and to measure switch costs, we conducted logistic and linear mixed effects models similar to those described above with trial type (effect coded: -1 = task repetition, 1= task switch), disease, drug state and session as predictors. All regression models included subject-specific random intercepts and random slopes for all within-subjects variables. RTs were log-transformed before being entered in the analysis to remove skew.

Given the differences in the SDMT performance between Parkinson’s patients and healthy controls (Table 1), we added participants’ z-scored SDMT score as an additional predictor in all our models. In cases in which the SDMT score produced a significant main effect or in which the effect of the SDMT score differed between experimental groups (i.e., an interaction effect involving the SDMT score and either disease or drug state), we report the coefficients of the regression model including this predictor. However, this was only the case for the regression model predicting choice RTs (see supplementary material). If there were no significant effects, we present only results from the simpler regression models (i.e., excluding the SDMT score).

## 3. Results

### 3.1 Dopaminergic medication reduces effort avoidance in Parkinson’s patients

As expected, overall, both healthy controls and Parkinson’s patients appeared to prefer the low-demand option over the high-demand option in the DST (*M* = 54.79%, *SD* = 13.53), indicating a general aversion to cognitive effort (Figure 1A). A one-sample t-test revealed that the higher proportion of low-demand choices was significantly different from 50% (*t*(61) = 2.96, *p* = .002). Patients OFF their dopaminergic medication exhibited the highest amount of low-demand choices (*M* = 57.11%, *SD* = 15.80, *t*(28) = 3.34, *p* = .011), whereas patients ON dopamine seemed to be more indifferent to task demand (*M* = 52.68%, *SD* = 11.77, *t*(35) = 1.36, *p* = .090). Control participants’ demand preferences fell in between the two patient groups (*M* = 55.14%, *SD* = 13.11, *t*(23) = 1.92, *p* = .034). A mixed-effects logistic regression revealed that patients avoided choosing the high-demand option significantly more often when they were OFF compared to when they were ON dopaminergic medications (main effect drug state: *p* = .005; for full results see Table 2). Contrary to our initial hypothesis, patients OFF dopamine were not significantly more effort-avoidant than healthy controls (main effect disease state: *p* = .432).

**Table 2.** Mixed-Effects Logistic Regression predicting low-demand choices in the free-choice blocks of the DST as a function of disease state, drug state, trial number and session. *Note*. *** *p* < .001, ** *p* < .01, * *p* < .05

| Predictor | *b* (*SE*) | *p* - value |
| --- | --- | --- |
| Intercept | 0.299 (0.113) | .008 ** |
| Disease state | -0.114 (0.145) | .432 |
| Drug state | -0.436 (0.154) | .005 ** |
| Trial number | 0.002 (0.004) | .653 |
| Session | 0.030 (0.108) | .780 |
| Disease state × trial number | 0.001 (0.006) | .823 |
| Drug state × trial number | 0.008 (0.003) | .003 ** |
| Disease state × session | -0.055 (0.143) | .700 |
| Drug state × session | -0.159 (0.140) | .255 |

**Fig. 1.**
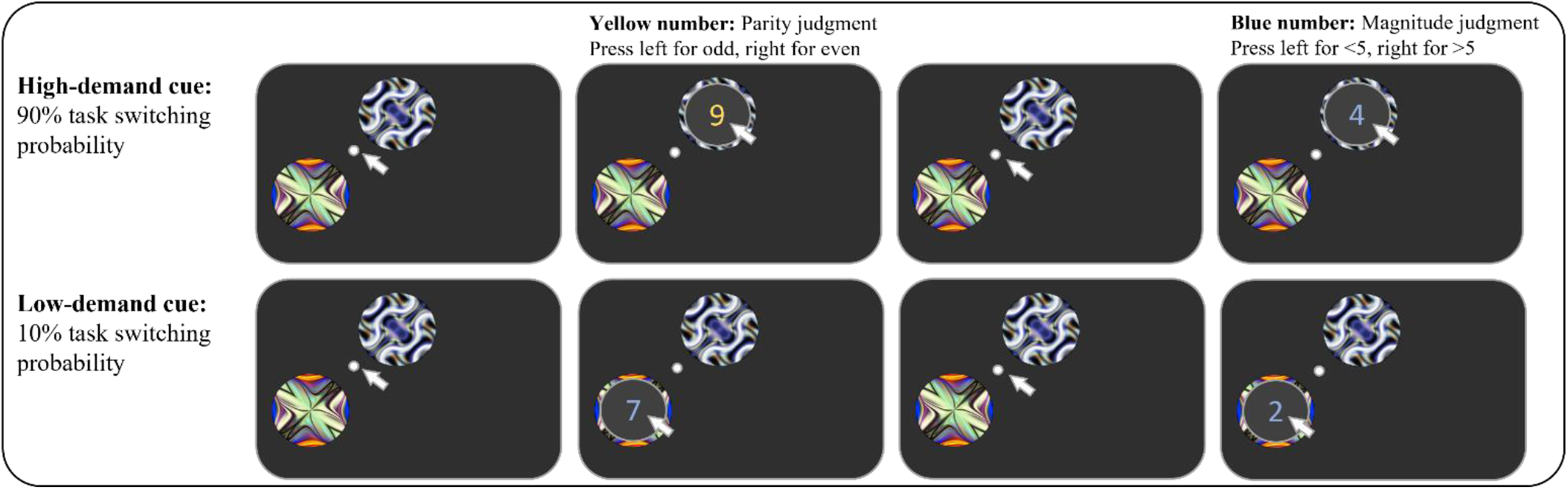
Demand Selection Task (DST). Participants choose to move the mouse cursor over one of two abstract color patches. The selected patch will then reveal a number (range: 1 – 4 and 6 – 9) that prompts participants to judge either its magnitude (i.e., smaller or larger than 5) or parity (i.e., whether it is odd or even) based on its color. Color patches differ in the probability with which the revealed numbers change their color from trial to trial and thus in the frequency participants have to switch between magnitude and parity judgment (high-demand patch: 90% switch rate, low-demand patch: 10% switch rate). Figure adapted from Bogdanov et al. (2021).

We next examined potential changes in demand preferences over time (Figure 2C). While preferences appeared to be relatively stable over the course of DST blocks for OFF patients (main effect trial number: *p* = .653) and controls (disease state × trial number interaction: *p* = .823), the proportion of low-demand choices of patients ON dopamine increased over time (drug state × trial number interaction: *p* = .003), indicating either slower learning of the difference in demand levels between the two options, or a weaker initial preference against effort exertion. There were no significant differences between sessions in either group (main effect session: *p* = .392, disease × session interaction: *p* = .700, drug state × session interaction: *p* = .255), indicating that there were no carry-over effects across testing days.

**Fig. 2.**
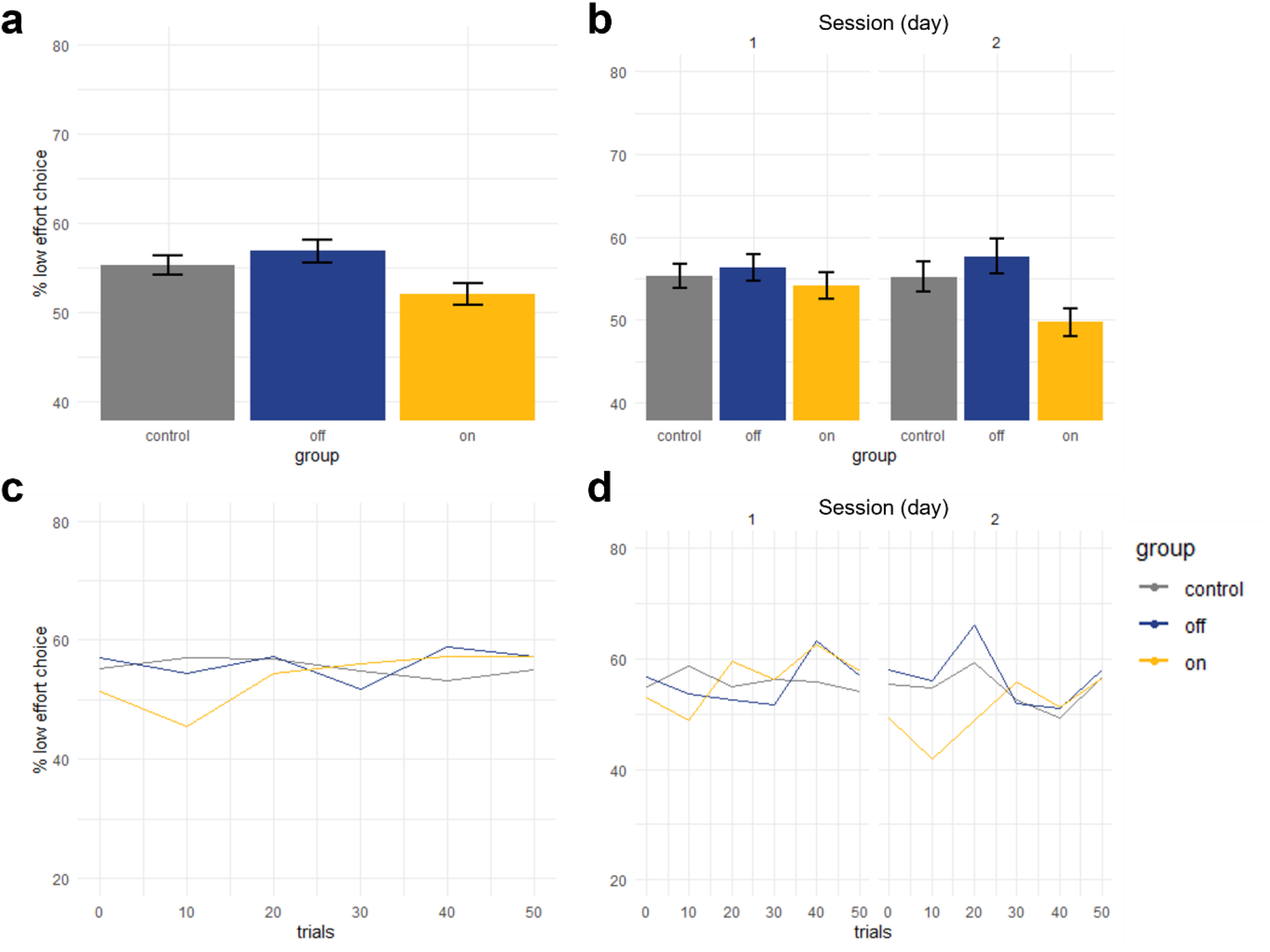
Choice behavior in the free-choice blocks of the DST. Overall, Parkinson’s patients showed a higher degree of effort avoidance when they were OFF dopaminergic medication compared to ON. Control participants’ proportion of low-demand choices was comparable to patients OFF dopamine (a). This pattern was found in both testing sessions (b). For patients OFF dopamine, the proportion of low-demand choices was particularly low at the beginning of a block and increased over the course of a block both overall (c) and in each session (d), whereas Parkinson’s patients ON dopamine and healthy controls where more effort avoidant early in the block and stayed consistent over time.

To rule out the possibility that disease or medication states altered participants’ ability to detect differences in demand levels in the DST, we calculated a separate mixed effects logistic regression to predict the proportion of correct choices in the forced-choice blocks, wherein participants were explicitly instructed to choose the high-or low-demand option (Gold et al., 2015; see Table 3 and Figure 3a). Results from these blocks suggest that participants were able to adapt their choice behavior to the instructions and that neither healthy controls (main effect disease state: *p* = .205) nor patients ON dopamine (main effect drug state: *p* = .752) differed significantly in their detection ability from patients OFF dopamine. We also found a significant increase in the proportion of correct choices in the instructed blocks over time when patients were OFF dopamine (main effect trial number: *p* <.001) and that the rate of this increase was higher compared to when patients were ON dopamine (drug state × trial number interaction: *p* <.001).

**Table 3.** Mixed-Effects Logistic Regression predicting instructed choices in the forced-choice blocks of the DST as a function of disease state, drug state, trial number, and session. *Note*. *** *p* < .001, ** *p* < .01, * *p* < .05

| Predictor | *b* (*SE*) | *p* - value |
| --- | --- | --- |
| Intercept | 0.094 (0.165) | .570 |
| Disease state | -0.059 (0.206) | .773 |
| Drug state | 0.088 (0.248) | .723 |
| Trial number | 0.051 (0.012) | <.001 *** |
| Session | 0.198 (0.147) | .180 |
| Disease state × trial number | -0.012 (0.019) | .512 |
| Drug state × trial number | -0.035 (0.008) | <.001 *** |
| Disease state × session | -0.189 (0.205) | .357 |
| Drug state × session | -0.158 (0.165) | .339 |

**Fig. 3.**
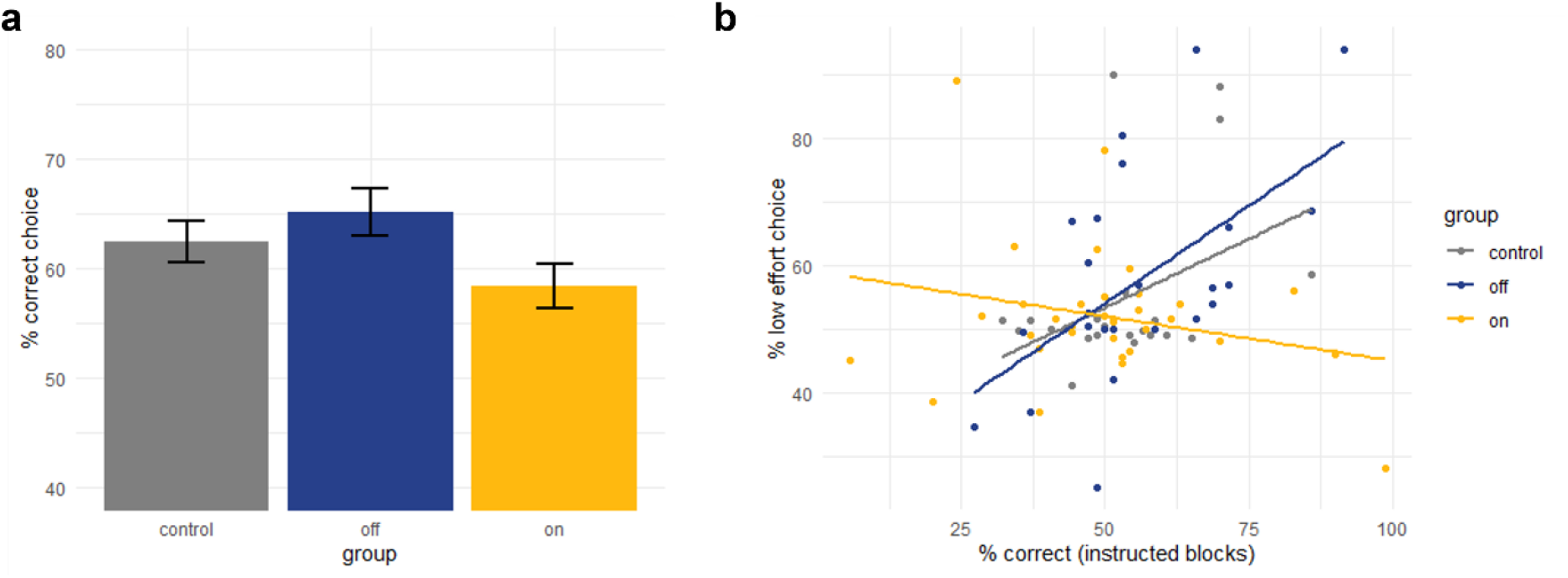
Detection performance in the forced-choice blocks of the DST. Participants in all groups were able to detect the differences in effort demand between the color patches and were able to adapt their choice behavior in accordance with the instructions (a). Better detection ability was related to more effort avoidant choice behavior for both healthy controls and Parkinson’s patients OFF dopamine, whereas for patients ON dopamine, the proportion of low-demand choices was independent of their ability to discriminate between the low- and high-demand color patches.

In order to further investigate how the individuals’ ability to detect the demand difference between the two options, we calculated a separate mixed effects logistic regression in which we added detection ability (i.e., z-scored percentage of correct choices in instructed blocks) as an additional main effect as well as its interactions with disease state and medication state (Table 4). Results of this exploratory analysis revealed that patients OFF dopamine who exhibited better detection ability in the instructed blocks were more likely to choose the low-demand patch in the free-choice blocks (main effect detection ability: *p* =.024). Similar behavior was observed in healthy controls (disease state × detection ability interaction: *p* = .676) but not in patients ON dopamine (drug state × detection ability interaction: *p* = .002) whose choices appeared to be unrelated to their ability to discriminate between the option’s demand levels (Figure 3b).

**Table 4.** Mixed-Effects Logistic Regression predicting low-demand choices in the DST from disease state, drug state, trial number, session, and detection ability. *Note*. Detection ability = z-scored proportion of choices according to instruction the forced-choice blocks of the DST. *** *p* < .001, ** *p* < .01, * *p* < .05

| Predictor | *b* (*SE*) | *p* - value |
| --- | --- | --- |
| Intercept | 0.256 (0.106) | .016 * |
| Disease state | -0.062 (0.149) | .676 |
| Drug state | -0.316 (0.118) | .007 ** |
| Trial number | -0.001 (0.004) | .948 |
| Session | 0.130 (0.103) | .210 |
| Detection ability | 0.320 (0.141) | .024 * |
| Disease state × trial number | 0.002 (0.005) | .780 |
| Drug state × trial number | 0.009 (0.003) | .002 ** |
| Disease state × session | -0.103 (0.129) | .427 |
| Drug state × session | -0.263 (0.135) | .051 |
| Disease state × detection ability | -0.169 (0.195) | .386 |
| Drug state × detection ability | -0.489 (0.157) | .002 ** |

Finally, in addition to participants’ choice preferences, we also analyzed choice reaction times (RTs) in the free-choice blocks of the DST, i.e., how long it took participants to move the mouse to the chosen option and click. Briefly, choice RTs in all groups decreased over the course of a block, with control participants being faster than patients both ON and OFF dopamine. Higher SDMT scores were associated with faster RTs in all groups. Full results can be found in the supplemental material (Table S1 and Figure S1).

### 3.2 Differences in choice behavior are not explained by task-switching performance

We also analyzed the effects of disease and medication state on performance in the task-switching portion of the DST (Table 5). Overall accuracy was very high in all groups (controls: *M* = 98.40%, *SD* = 2.14; PD OFF: *M* = 94.40%, *SD* = 9.40; PD ON: *M* = 94.49%, *SD* = 9.16) and did not differ between OFF patients and controls (main effect of disease state: *p* = .943) or ON patients (main effect drug state: *p* = .081; see Figure 4a). Similarly, accuracy was higher on task repetition trials (i.e., trials in which the colored number signalled the same type of judgment as in the trial before) compared to task switch trials in all experimental groups (main effect trial type: *p* = <.001; disease state × trial type interaction: *p* = .978; drug state × trial type interaction: *p* = .531).

**Table 5.** Results of a Mixed-Effects Logistic Regression predicting task-switching accuracy and a Mixed-Effects Linear Regression predicting task-switching reaction times in the DST from disease state, drug state, trial type and session. *Note*. *** *p* < .001, ** *p* < .01, * *p* < .05

| Predictor | Accuracy |  |  | Reaction times |  |  |
| --- | --- | --- | --- | --- | --- | --- |
|  | *b* (SE) | *p* - value |  | *b* (SE) | *p* - value |  |
| Intercept | 0.294 (0.132) | .025 | * | 0.186 (0.023) | <.001 | *** |
| Disease state | -0.014 (0.188) | .943 |  | -0.031 (0.035) | .372 |  |
| Drug state | -0.229 (0.131) | .081 |  | 0.006 (0.020) | .766 |  |
| Trial type | -2.149 (0.047) | <.001 | *** | 0.030 (0.007) | <.001 | *** |
| Session | -0.042 (0.132) | .750 |  | -0.022 (0.023) | .338 |  |
| Disease state × trial type | 0.002 (0.061) | .978 | | 0.004 (0.010) | .705 | |
| Drug state × trial type | -0.039 (0.062) | .531 | | 0.001 (0.006) | .972 | |
| Disease state × session | 0.049 (0.158) | .757 | | -0.012 (0.027) | .673 | |
| Drug state × session | -0.089 (0.212) | .673 | | -0.024 (0.040) | .547 | |

**Fig. 4.**
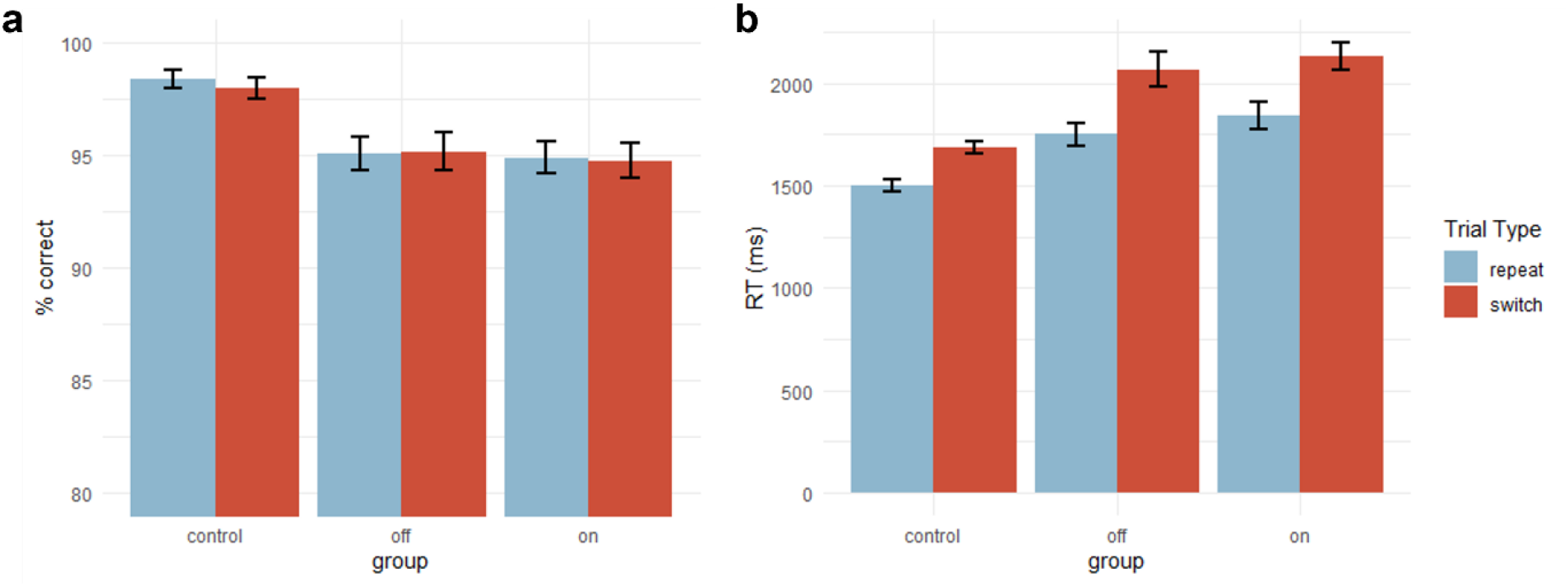
Task-switching performance in the free-choice blocks of the DST. All participants demonstrated high accuracy in the number judgment part of the DST, with overall more correct choices in repeat compared to switch trials (a). Reaction times (b) were slower for switch compared to repeat trials. Switch costs were similar across all groups.

Task-switching reaction times in the DST followed a similar pattern (Figure 4b). Overall, RTs do not differ significantly between OFF patients and controls (main effect disease state: *p* = .372) or between patients OFF and ON dopamine (main effect drug state: *p*= .766). Patients OFF dopamine exhibited typical RT switch costs (Monsell, 2003), i.e., longer RTs in switch compared to repeat trials (main effect trial type: *p* < .001). Again, the magnitude of these switch costs were not statistically different from those observed in controls (disease state × trial type interaction: *p* = .705) and in ON patients (drug state × trial type interaction: *p* = .972).

In short, the absence of performance differences (either in accuracy or RTs) in the task-switching paradigm within the DST between conditions suggests that the gap in effort avoidance between ON and OFF dopamine patients is unlikely to result from more general modulations of the patients’ cognitive capabilities.

### 3.3 Individual differences in switch costs do not predict effort avoidance in Parkinson’s disease

We assumed that participants with larger RT switch costs—that is, a larger RT cost incurred by switching, versus repeating tasks—may have experienced the task-switching paradigm in the DST as particularly effortful (Bogdanov et al., 2021; Kool et al., 2010). To explore whether the magnitude of participants switch costs in the number judgment task, either in the DST (across all trials) or in the baseline task-switching paradigm, may relate to preferences for the low-demand option in the DST, we calculated two additional mixed-effects logistic regressions predicting low-demand choice, in which we added z-scored RT task switch costs (DST or baseline) as a main effect as well as their interactions with disease state and drug state (Table 6). These analyses, visualized in Figure 5, suggest that neither RT switch costs estimated in DST nor in the baseline task-switching paradigm predicted choice behavior for Parkinson’s patients OFF dopamine (main effects switch costs: *p* = .813 and *p* =.462, respectively) or ON dopamine (drug state × switch costs interactions: *p* = .977 and *p* =.904, respectively). We did, however observe marginally significant predictive relationships between the magnitude of switch costs measured in the DST as well as at baseline and effort-avoidant choice behavior in healthy controls (disease state × detection ability interactions: *p* =.050 and *p* = .069, respectively), suggesting that these participants were able to adjust their choice preferences in line with the individual effort costs of task-switching (Bogdanov et al., 2021; Liu & Yeung, 2020; Monsell & Mizon, 2006), whereas this link was weakened in Parkinson’s patients.

**Table 6.**
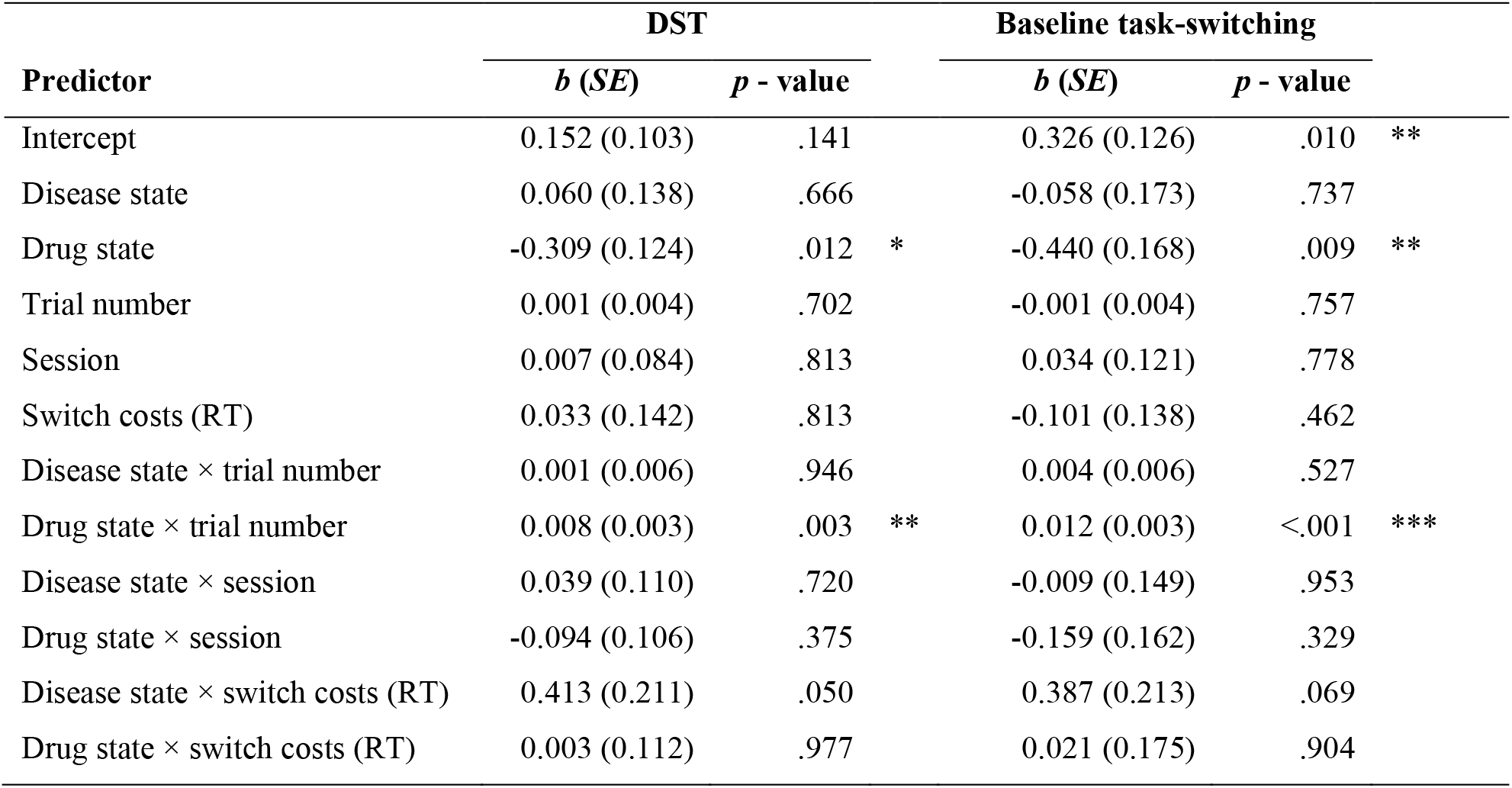
Results of a Mixed-Effects Logistic Regression predicting low-demand choices in the free-choice blocks of the DST as a function of disease state, drug state, trial number, session and RT switch costs in both the DST and the baseline task-switching task.*Note*. *** *p* < .001, ** *p* < .01, * *p* < .05

**Fig. 5.**
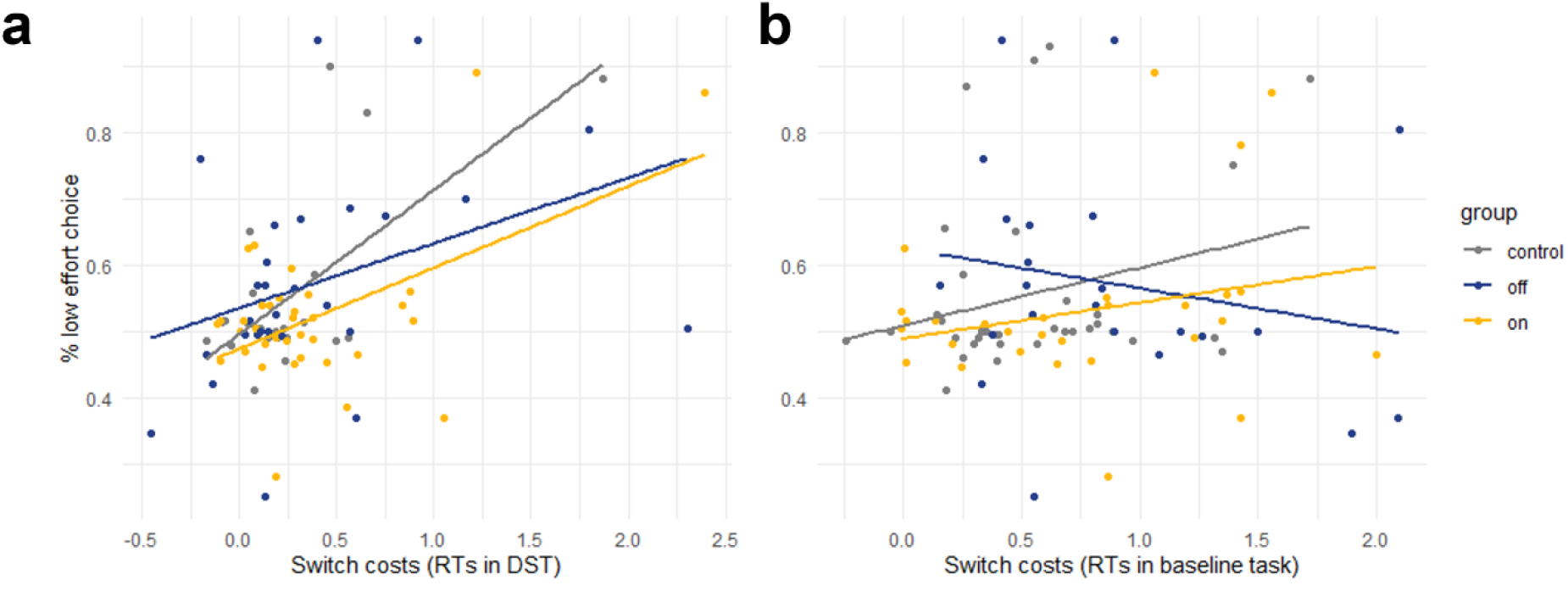
RT switch costs in relation to choice behavior in the DST. Descriptively, participants with larger RT switch costs in the number judgment part of the DST showed stronger effort avoidant choice behavior (a). A similar descriptive pattern was seen for control participants and patients ON dopamine, but not for patients OFF dopamine, with respect to RT switch costs in the baseline task switching paradigm (b).

In addition, we also analyzed whether the individuals’ degree of apathy (measured by the AES) or depression (measured by the GDS) predicted choice behavior in the DST. In short, we did not find a significant effect of either measure on participants’ effort avoidance. Full results from these analyses can be found in the supplement (Table S2 and Figure S2).

## 4. Discussion

The strategic allocation of our limited cognitive resources to pursue a desired goal represents a key function of successful behavioral adaptation in everyday life (Chong et al., 2017; Kool & Botvinick, 2018; Shenhav et al., 2017; Westbrook & Braver, 2015). While previous research heavily implies a role of dopamine specifically in increasing the willingness to deploy cognitive effort, much of this work has focused on investigating the postulated cost-benefit trade-off between effort and expected reward, suggesting that dopamine leads to increased sensitivity to the benefits conferred by effort deployment relative its costs (McGuigan et al., 2019; Salamone et al., 2016; Watson et al., 1988; Westbrook et al., 2020). Given dopamine’s role in increasing the capacity for cognitive control (Westbrook & Braver, 2016), it is plausible that dopamine would also cause a reduction in the sensitivity to the costs of cognitive effort even in the absence of any clear incentives, but this remains largely untested (Froböse et al., 2018). Using a well-established demand-selection paradigm where only the level of effort, but not reward, is manipulated (Bogdanov et al., 2021; Gold et al., 2015; Kool et al., 2010; Patzelt et al., 2019), we show that dopaminergic medication decreases effort-avoidant behavior in Parkinson’s patients. More specifically, both healthy controls and Parkinson’s patients OFF dopamine displayed the expected preference for low-demand options in the DST whereas patients ON dopamine chose the low- and high-demand options equally often. Importantly, the increased willingness to exert effort observed in the patients ON could not be attributed to differences in the participants’ ability to detect the harder task nor to medication-induced changes in performance on the cognitive control task, making it unlikely that the reduced demand avoidance in patients ON could be attributed to medication-related performance improvements.

Current accounts of cognitive effort-based decision-making emphasize the cost-benefit trade-off between the costs of effort exertion and the potential rewards for successful task completion (Kool & Botvinick, 2018; Shenhav et al., 2017; Westbrook & Braver, 2015). Indeed, a growing body of literature has demonstrated that the prospect of cognitive effort exertion considerably reduces (or discounts) the subjective value, and thus the motivational draw, of a potential outcome, reducing the likelihood of an individual to engage in the associated behavior (Apps et al., 2015; Bogdanov et al., In press; Chong et al., 2017; Massar et al., 2015; Vogel et al., 2020; Westbrook et al., 2013). Given the well-established involvement of dopamine in reward processing (Aarts et al., 2012; Berridge & Robinson, 1998; Buckholtz et al., 2010; Schultz, 2013; Sharp et al., 2015, 2020), effects of dopaminergic medication on effort allocation in studies that manipulate both could thus in principle arise from modulations of reward instead of effort-related calculations (Chong et al., 2015; McGuigan et al., 2019; Michely et al., 2020). Our findings, on the other hand, demonstrate that dopamine modulates an individual’s willingness to exert cognitive effort even when exerting more effort does not confer greater rewards, as in the DST, thus arguing for a more direct and specific effect of dopamine on participants’ sensitivity to effort costs.

Two possible mechanisms could explain the reduced demand avoidance we observed in patients ON compared to patients OFF dopaminergic medications: dopamine replacement could lead to a decrease in the feelings of subjective aversiveness associated with cognitive effort exertion (Kurzban, 2016; Vogel et al., 2020), or dopamine could increase the availability of cognitive resources, thereby reducing the costs associated with effort exertion. Although the DST is not designed to disentangle these two possible mechanisms, the fact that there were no differences in task performance between the patients ON and OFF (both accuracy and reaction times in the task-switching portion of the DST were comparable across medication states), suggests that the reduced effort avoidance in Parkinson’s patients ON medication was not simply due to enhanced capacity in this group. We also included two forced-choice blocks where patients were either instructed to choose the ‘easy’ option or to choose the ‘hard’ option. These were added to ensure that differences in effort avoidance could not be attributed to differences in the ability to detect cognitive demand levels, as has been shown to be the case in schizophrenia (Gold et al., 2015). We found no group differences in the ability to accurately identify the effort demand level of the two options. Moreover, as expected, better detection ability was associated with a higher tendency to avoid the high demand option in healthy controls and patients OFF dopaminergic medications. In contrast, patients ON dopamine did not show a relationship between detection ability and effort avoidance. In other words, the patients ON medication chose to exert more cognitive effort than patients OFF despite similar awareness of the differences in task demand between the two color patches.

Interestingly, the difference in effort avoidance between ON and OFF patients was larger at the beginning of an experimental block, suggesting that the initially decreased effort sensitivity under medication may wane over time. Alternatively, this result could suggest that Parkinson’s patients on dopamine required more time to learn about the differences in task demand associated with both cues in the DST. Indeed, there is ample evidence for altered learning processes in patients with Parkinson’s disease (Pascual-Leone et al., 1993; Sharp et al., 2015, 2020), although this may be specific to learning about rewards instead of effort costs (Skvortsova et al., 2017). While learning is often thought to be impaired in Parkinson’s patients and to be improved by dopamine replacement, some evidence suggests that dopaminergic medication can be detrimental to learning processes that depend on brain regions not yet affected by the disease (Graef et al., 2010). We cannot conclusively distinguish between these possibilities in our study but given that patients performed equally well in the forced-choice blocks of the DST both ON and OFF medication, it seems less likely that the smaller proportion of low-demand choices at the beginning of a block observed in patients ON dopamine simply reflects impaired learning in this group.

Our findings also fit well with earlier work in both humans in animals demonstrating that pharmacologically elevating dopamine levels can increase an individual’s willingness to engage in effort behavior (Bardgett et al., 2009; Floresco et al., 2008; Le Bouc et al., 2016; Michely et al., 2020). Many of these studies, however, rely on manipulating physical demand to investigate the role of dopamine in motivating effort exertion. Although it has been proposed that physical and cognitive effort may be distinct to a degree and that dopamine may primarily affect decisions about physical effort (Hosking et al., 2015), this domain-specific view has been challenged by more recent findings. For example, patients with Parkinson’s disease show more pronounced effort discounting in a visual attention-based task under dopamine withdrawal compared to when they are ON dopaminergic medication (McGuigan et al., 2019). Moreover, administration of methylphenidate, which increases dopamine and noradrenaline levels in the brain, increases participants’ motivation to expend more effort in working memory tasks (Froböse et al., 2018; Hofmans et al., 2020; Westbrook et al., 2020). Our results lend further support in favor of a domain general role of dopamine in motivating behavior and overcoming effort costs (Westbrook & Braver, 2016).

Contrary to our expectations, patients OFF dopamine, despite displaying the highest proportion of low-demand choices in the DST among the three groups, were not significantly more effort avoidant than healthy controls. This is surprising given that previous work demonstrated that reduced dopamine levels decrease motivation to exert effort for reward. For example, dopamine depleted rats are less inclined to climb over barriers or to repeatedly press levers for larger rewards compared to control rats (Denk et al., 2005; Floresco et al., 2008; Salamone et al., 2007). Similarly, Parkinson’s patients OFF dopamine have been shown to prefer to receive lower compared to higher rewards to avoid physical effort exertion in a grip-strength task (Chong et al., 2015; Le Heron et al., 2018). A possible explanation for the similar degree of effort avoidance in OFF patients and controls may be that the patients in our study represent a high functioning sub-sample of the broader Parkinson’s population, who were intrinsically motivated and physically able to participate in lab research. Patients and controls did not differ in general cognitive ability (measured by the MoCA), depressive symptoms, or symptoms of apathy, which may also explain why there was no relationship between these measures and the individuals’ effort sensitivity that has been reported in other studies (McGuigan et al., 2019). Another possibility is that aging-associated decline in midbrain dopaminergic function could result in changes to cognitive effort sensitivity (Kaasinen & Rinne, 2002). Indeed, previous studies of healthy older adults measuring dopamine-related cognitive processes have shown that dopamine replacement can restore impairments in a way that is similar to its effects in Parkinson’s disease even though the dopaminergic loss associated with aging is much less substantial than that seen in Parkinson’s disease (Chowdhury et al., 2012, 2013).

It should also be noted that the proportion of low-demand choices in our study is somewhat lower than in prior work using the DST (Bogdanov et al., 2021; Gold et al., 2015; Kool et al., 2010). This may have been a result of the particular design used in our experiment. More specifically, while some other studies kept the color patches and their associated task-switching frequencies constant across all blocks, patches in our experiment changed at the start of every new blocks and participants had to explore both options anew in each block. Given that there were only 50 trials per block, it is difficult to achieve high percentages of low-demand choices by the end the block. Based on reports that participants are able to develop stable choice preferences as early as 35 trials into the task (Gold et al., 2015), we aimed to keep blocks short to not overwhelm our patients and older controls. Indeed, as evidenced by participants’ performance in the forced-choice blocks, the ability to differentiate the high-demand and low-demand options emerged relatively quickly. However, we cannot dismiss the possibility that longer blocks or the use of a consistent set of color patches across all blocks would have allowed for the development of more pronounced effort-avoidant choice behavior, and potentially resulted in differences between patients OFF and healthy controls. Future research might thus implement these changes to the protocol to maximize statistical power.

Our analyses examining the relationship between individual differences in task switch costs (in both the DST and the baseline task switching paradigm) and choice behavior in the DST revealed that, descriptively, healthy controls who exhibited larger switch costs, i.e., who showed less cognitive control capacity, were more likely to choose the low-demand option. In contrast, there was no relationship between switch cost magnitude and choice behavior in Parkinson’s patients, irrespective of medication status. Although these exploratory findings should be interpreted with caution and require replication, they may suggest that Parkinson’s disease diminishes the participants’ ability to strategically adjust their demand preferences according to their cognitive abilities. Our observations are in line with earlier findings in patients with Schizophrenia, who have been shown to demonstrate a similar deficit in monitoring their subjective costs of effort allocation (Gold et al., 2015). Interestingly, we also did not find dopamine-related modulations of response times in the task-switching part of the DST. Given that tonic dopamine levels have been associated with increased response vigor (Beierholm et al., 2013; Niv et al., 2007; Zénon et al., 2016), one could have expected patients

ON medication to show a general speeding up of responses to the parity and magnitude judgments, potentially to the expense of accuracy. However, we did not observe such a pattern. In fact, participants in all groups were highly accurate in their number judgment responses. This may in part be explained by the fact that older adults generally value correct over fast responses (Rabbitt, 1979; Starns & Ratcliff, 2010), possibly overshadowing an increase in speed due to dopamine.

In conclusion, our study provides evidence that dopamine may play a direct role in increasing the motivation to exert cognitive effort, above and beyond its role in increasing the sensitivity to the benefits of effort exertion. The effect of dopaminergic medications on the willingness to exert effort could not be explained by differences in cognitive control capacity, nor by differences in the ability to detect effort demands in the DST. Taken together, this suggests that dopamine may specifically modulate participants’ sensitivity to or calculation of cognitive effort costs. Our results lend further support to the hypothesis that dopamine plays an important and domain-general role in guiding the strategic allocation of both physical and mental resources to adjust behavior. Although more work is necessary to fully characterize the underlying mechanisms of how dopamine modulates motivational processes, our findings may be a valuable contribution to the collective effort to improve our understanding and treatment options of severe reductions in motivation that are common in Parkinson’s disease and, given the potential transdiagnostic properties of cognitive effort-based decision-making impairments (Patzelt et al., 2019), in many other psychiatric and neurologic conditions.

## Supporting information

supplemental_results

## Data Availability

All data produced in the present study are available upon reasonable request to the authors

## Authorship contributions

### Mario Bogdanov

Formal analysis, Writing – Original Draft, Visualization. **Sophia LoParco:** Methodology, Data Collection, Data Curation **A. Ross Otto:** Conceptualization, Methodology, Resources, Writing – Review & Editing, Supervision. **Madeleine Sharp:** Conceptualization, Methodology, Resources, Data Curation, Writing – Review & Editing, Supervision, Project Administration, Funding Acquisition.

## Acknowledgements

The authors thank Léah Suissa-Rocheleau, Elsie Yan, Soraya Lahlou and Matthew Pilgrim for their assistance with data collection. This project was funded by the Parkinson Foundation and the Fonds de Recherche du Québec - Santé to M.S., the NSERC Discovery Grant and the New Researchers Startup Grant from the Fonds de Recherche du Québec - Nature et Technologies to A.R.O., and by a postdoctoral fellowship by the German Research Foundation (DFG) to M.B.

## References

Aarts, E., Helmich, R. C., Janssen, M. J., Oyen, W. J., Bloem, B. R., & Cools, R. (2012). Aberrant reward processing in Parkinson’s disease is associated with dopamine cell loss. Neuroimage, 59(4), 3339–3346.

Apps, M. A., Grima, L. L., Manohar, S., & Husain, M. (2015). The role of cognitive effort in subjective reward devaluation and risky decision-making. Scientific Reports, 5, 16880.

Bardgett, M. E., Depenbrock, M., Downs, N., Points, M., & Green, L. (2009). Dopamine modulates effort-based decision making in rats. Behavioral Neuroscience, 123(2), 242.

Bates, D., Mächler, M., Bolker, B., & Walker, S. (2014). Fitting linear mixed-effects models using lme4. ArXiv Preprint 1406.5823.

Beierholm, U., Guitart-Masip, M., Economides, M., Chowdhury, R., Düzel, E., Dolan, R., & Dayan, P. (2013). Dopamine modulates reward-related vigor. Neuropsychopharmacology, 38(8), 1495–1503.

Berridge, K. C., & Robinson, T. E. (1998). What is the role of dopamine in reward: Hedonic impact, reward learning, or incentive salience? Brain Research Reviews, 28(3), 309–369.

Bódi, N., Kéri, S., Nagy, H., Moustafa, A., Myers, C. E., Daw, N., Dibó, G., Takats, A., Bereczki, D., & Gluck, M. A. (2009). Reward-learning and the novelty-seeking personality: A between-and within-subjects study of the effects of dopamine agonists on young Parkinson’s patients. Brain, 132(9), 2385–2395.

Bogdanov, M., Nitschke, J. P., LoParco, S., Bartz, J. A., & Otto, A. R. (2021). Acute Psychosocial Stress Increases Cognitive-Effort Avoidance. Psychological Science, 09567976211005465.

Bogdanov, M., Renault, H., LoParco, S., Weinberg, A., & Otto, A. R. (n.d.). Cognitive effort exertion enhances electrophysiological responses to rewarding outcomes.

Brown, D. R., Richardson, S. P., & Cavanagh, J. F. (2020). An EEG marker of reward processing is diminished in Parkinson’s disease. Brain Research, 1727, 146541.

Buckholtz, J. W., Treadway, M. T., Cowan, R. L., Woodward, N. D., Benning, S. D., Li, R., Ansari, M. S., Baldwin, R. M., Schwartzman, A. N., & Shelby, E. S. (2010). Mesolimbic dopamine reward system hypersensitivity in individuals with psychopathic traits. Nature Neuroscience, 13(4), 419–421.

Chong, T. T.-J., Apps, M., Giehl, K., Sillence, A., Grima, L. L., & Husain, M. (2017). Neurocomputational mechanisms underlying subjective valuation of effort costs. PLoS Biology, 15(2), e1002598.

Chong, T. T.-J., Bonnelle, V., Manohar, S., Veromann, K.-R., Muhammed, K., Tofaris, G. K., Hu, M., & Husain, M. (2015). Dopamine enhances willingness to exert effort for reward in Parkinson’s disease. Cortex, 69, 40–46.

Chowdhury, R., Guitart-Masip, M., Bunzeck, N., Dolan, R. J., & Düzel, E. (2012). Dopamine modulates episodic memory persistence in old age. Journal of Neuroscience, 32(41), 14193–14204.

Chowdhury, R., Guitart-Masip, M., Lambert, C., Dayan, P., Huys, Q., Düzel, E., & Dolan, R. J. (2013). Dopamine restores reward prediction errors in old age. Nature Neuroscience, 16(5), 648–653.

Cocker, P. J., Hosking, J. G., Benoit, J., & Winstanley, C. A. (2012). Sensitivity to cognitive effort mediates psychostimulant effects on a novel rodent cost/benefit decision-making task. Neuropsychopharmacology, 37(8), 1825–1837.

Cools, R. (2016). The costs and benefits of brain dopamine for cognitive control. Wiley Interdisciplinary Reviews: Cognitive Science, 7(5), 317–329.

da Silva Castanheira, K., LoParco, S., & Otto, A. R. (2021). Task-evoked pupillary responses track effort exertion: Evidence from task-switching. Cognitive, Affective, & Behavioral Neuroscience, 21(3), 592–606.

den Brok, M. G., van Dalen, J. W., van Gool, W. A., Moll van Charante, E. P., de Bie, R. M., & Richard, E. (2015). Apathy in Parkinson’s disease: A systematic review and meta-analysis. Movement Disorders, 30(6), 759–769.

Denk, F., Walton, M. E., Jennings, K. A., Sharp, T., Rushworth, M. F. S., & Bannerman, D. M. (2005). Differential involvement of serotonin and dopamine systems in cost-benefit decisions about delay or effort. Psychopharmacology, 179(3), 587–596.

Dreisbach, G., & Haider, H. (2006). Preparatory adjustment of cognitive control in the task switching paradigm. Psychonomic Bulletin & Review, 13(2), 334–338.

Floresco, S. B., Maric, T., & Ghods-Sharifi, S. (2008). Dopaminergic and glutamatergic regulation of effort-and delay-based decision making. Neuropsychopharmacology, 33(8), 1966–1979.

Froböse, M. I., Swart, J. C., Cook, J. L., Geurts, D. E., Den Ouden, H. E., & Cools, R. (2018). Catecholaminergic modulation of the avoidance of cognitive control. Journal of Experimental Psychology: General, 147(12), 1763.

Gelman, A., & Hill, J. (2006). Data analysis using regression and multilevel/hierarchical models. Cambridge university press.

Gold, J. M., Kool, W., Botvinick, M. M., Hubzin, L., August, S., & Waltz, J. A. (2015). Cognitive effort avoidance and detection in people with schizophrenia. Cognitive, Affective, & Behavioral Neuroscience, 15(1), 145–154.

Graef, S., Biele, G., Krugel, L. K., Marzinzik, F., Wahl, M., Wotka, J., Klostermann, F., & Heekeren, H. R. (2010). Differential influence of levodopa on reward-based learning in Parkinson’s disease. Frontiers in Human Neuroscience, 4, 169.

Hofmans, L., Papadopetraki, D., van den Bosch, R., Määttä, J. I., Froböse, M. I., Zandbelt, B. B., Westbrook, A., Verkes, R.-J., & Cools, R. (2020). Methylphenidate boosts choices of mental labor over leisure depending on striatal dopamine synthesis capacity. Neuropsychopharmacology, 45(13), 2170–2179.

Hosking, J. G., Floresco, S. B., & Winstanley, C. A. (2015). Dopamine antagonism decreases willingness to expend physical, but not cognitive, effort: A comparison of two rodent cost/benefit decision-making tasks. Neuropsychopharmacology, 40(4), 1005–1015.

Hull, C. L. (1943). Principles of behavior: An introduction to behavior theory.

Husain, M., & Roiser, J. P. (2018). Neuroscience of apathy and anhedonia: A transdiagnostic approach. Nature Reviews Neuroscience, 19(8), 470.

Inzlicht, M., Shenhav, A., & Olivola, C. Y. (2018). The effort paradox: Effort is both costly and valued. Trends in Cognitive Sciences, 22(4), 337–349.

Kaasinen, V., & Rinne, J. O. (2002). Functional imaging studies of dopamine system and cognition in normal aging and Parkinson’s disease. Neuroscience & Biobehavioral Reviews, 26(7), 785–793.

Kleiner, M., Brainard, D., & Pelli, D. (2007). What’s new in Psychtoolbox-3?

Kool, W., & Botvinick, M. (2018). Mental labour. Nature Human Behaviour, 2(12), 899–908.

Kool, W., McGuire, J. T., Rosen, Z. B., & Botvinick, M. M. (2010). Decision making and the avoidance of cognitive demand. Journal of Experimental Psychology: General, 139(4), 665.

Kurzban, R. (2016). The sense of effort. Current Opinion in Psychology, 7, 67–70.

Kurzban, R., Duckworth, A., Kable, J. W., & Myers, J. (2013). An opportunity cost model of subjective effort and task performance. Behavioral and Brain Sciences, 36(6), 661–679.

Le Bouc, R., Rigoux, L., Schmidt, L., Degos, B., Welter, M.-L., Vidailhet, M., Daunizeau, J., & Pessiglione, M. (2016). Computational dissection of dopamine motor and motivational functions in humans. Journal of Neuroscience, 36(25), 6623–6633.

Le Heron, C., Plant, O., Manohar, S., Ang, Y.-S., Jackson, M., Lennox, G., Hu, M. T., & Husain, M. (2018). Distinct effects of apathy and dopamine on effort-based decision-making in Parkinson’s disease. Brain, 141(5), 1455–1469.

Lemke, M. R., Brecht, H. M., Koester, J., Kraus, P. H., & Reichmann, H. (2005). Anhedonia, depression, and motor functioning in Parkinson’s disease during treatment with pramipexole. The Journal of Neuropsychiatry and Clinical Neurosciences, 17(2), 214–220.

Liu, C., & Yeung, N. (2020). Dissociating expectancy-based and experience-based control in task switching. Journal of Experimental Psychology: Human Perception and Performance, 46(2), 131.

Manohar, S. G., Chong, T. T.-J., Apps, M. A., Batla, A., Stamelou, M., Jarman, P. R., Bhatia, K. P., & Husain, M. (2015). Reward pays the cost of noise reduction in motor and cognitive control. Current Biology, 25(13), 1707–1716.

Massar, S. A., Libedinsky, C., Weiyan, C., Huettel, S. A., & Chee, M. W. (2015). Separate and overlapping brain areas encode subjective value during delay and effort discounting. Neuroimage, 120, 104–113.

Mazzoni, P., Hristova, A., & Krakauer, J. W. (2007). Why don’t we move faster? Parkinson’s disease, movement vigor, and implicit motivation. Journal of Neuroscience, 27(27), 7105–7116.

McGuigan, S., Zhou, S.-H., Brosnan, M. B., Thyagarajan, D., Bellgrove, M. A., & Chong, T. T. (2019). Dopamine restores cognitive motivation in Parkinson’s disease. Brain, 142(3), 719–732.

Michely, J., Viswanathan, S., Hauser, T. U., Delker, L., Dolan, R. J., & Grefkes, C. (2020). The role of dopamine in dynamic effort-reward integration. Neuropsychopharmacology, 45(9), 1448–1453.

Monsell, S. (2003). Task switching. Trends in Cognitive Sciences, 7(3), 134–140.

Monsell, S., & Mizon, G. A. (2006). Can the task-cuing paradigm measure an endogenous task-set reconfiguration process? Journal of Experimental Psychology: Human Perception and Performance, 32(3), 493.

Niv, Y., Daw, N. D., Joel, D., & Dayan, P. (2007). Tonic dopamine: Opportunity costs and the control of response vigor. Psychopharmacology, 191(3), 507–520.

Otto, A. R., & Daw, N. D. (2019). The opportunity cost of time modulates cognitive effort. Neuropsychologia, 123, 92–105.

Pascual-Leone, A., Grafman, J., Clark, K., Stewart, M., Massaquoi, S., Lou, J., & Hallett, M. (1993). Procedural learning in Parkinson’s disease and cerebellar degeneration. Annals of Neurology: Official Journal of the American Neurological Association and the Child Neurology Society, 34(4), 594–602.

Pasquereau, B., & Turner, R. S. (2013). Limited encoding of effort by dopamine neurons in a cost–benefit trade-off task. Journal of Neuroscience, 33(19), 8288–8300.

Patzelt, E. H., Kool, W., Millner, A. J., & Gershman, S. J. (2019). The transdiagnostic structure of mental effort avoidance. Scientific Reports, 9(1), 1–10.

Pilgrim, M. J., Ou, Z.-Y. A., & Sharp, M. (2021). Exploring reward-related attention selectivity deficits in Parkinson’s disease. Scientific Reports, 11(1), 1–11.

R Core Team. (2020). R: A language and environment for statistical computing (3.6. 2 (2019-12-12)). The R Foundation for Statistical Computing.

Rabbitt, P. (1979). How old and young subjects monitor and control responses for accuracy and speed. British Journal of Psychology, 70(2), 305–311.

Salamone, J. D., Correa, M., Farrar, A. M., Nunes, E. J., & Pardo, M. (2009). Dopamine, behavioral economics, and effort. Frontiers in Behavioral Neuroscience, 3, 13.

Salamone, J. D., Correa, M., Farrar, A., & Mingote, S. M. (2007). Effort-related functions of nucleus accumbens dopamine and associated forebrain circuits. Psychopharmacology, 191(3), 461–482.

Salamone, J. D., Correa, M., Yohn, S., Cruz, L. L., San Miguel, N., & Alatorre, L. (2016). The pharmacology of effort-related choice behavior: Dopamine, depression, and individual differences. Behavioural Processes, 127, 3–17.

Schott, B. H., Niehaus, L., Wittmann, B. C., Schütze, H., Seidenbecher, C. I., Heinze, H.-J., & Düzel, E. (2007). Ageing and early-stage Parkinson’s disease affect separable neural mechanisms of mesolimbic reward processing. Brain, 130(9), 2412–2424.

Schultz, W. (2013). Updating dopamine reward signals. Current Opinion in Neurobiology, 23(2), 229–238.

Sharp, M. E., Duncan, K., Foerde, K., & Shohamy, D. (2020). Dopamine is associated with prioritization of reward-associated memories in Parkinson’s disease. Brain, 143(8), 2519–2531.

Sharp, M. E., Foerde, K., Daw, N. D., & Shohamy, D. (2015). Dopamine selectively remediates ‘model-based’reward learning: A computational approach. Brain, 139(2), 355–364.

Shenhav, A., Musslick, S., Lieder, F., Kool, W., Griffiths, T. L., Cohen, J. D., & Botvinick, M. M. (2017). Toward a rational and mechanistic account of mental effort. Annual Review of Neuroscience, 40, 99–124.

Skvortsova, V., Degos, B., Welter, M.-L., Vidailhet, M., & Pessiglione, M. (2017). A selective role for dopamine in learning to maximize reward but not to minimize effort: Evidence from patients with Parkinson’s disease. Journal of Neuroscience, 37(25), 6087–6097.

Starns, J. J., & Ratcliff, R. (2010). The effects of aging on the speed–accuracy compromise: Boundary optimality in the diffusion model. Psychology and Aging, 25(2), 377.

Tanaka, S., Taylor, J. E., & Sakagami, M. (2021). The effect of effort on reward prediction error signals in midbrain dopamine neurons. Current Opinion in Behavioral Sciences, 41, 152–159.

Timmer, M. H., Aarts, E., Esselink, R. A., & Cools, R. (2018). Enhanced motivation of cognitive control in Parkinson’s disease. European Journal of Neuroscience, 48(6), 2374–2384.

Treadway, M. T., Bossaller, N. A., Shelton, R. C., & Zald, D. H. (2012). Effort-based decision-making in major depressive disorder: A translational model of motivational anhedonia. Journal of Abnormal Psychology, 121(3), 553.

Treadway, M. T., Buckholtz, J. W., Cowan, R. L., Woodward, N. D., Li, R., Ansari, M. S., Baldwin, R. M., Schwartzman, A. N., Kessler, R. M., & Zald, D. H. (2012). Dopaminergic mechanisms of individual differences in human effort-based decision-making. Journal of Neuroscience, 32(18), 6170–6176.

Varazzani, C., San-Galli, A., Gilardeau, S., & Bouret, S. (2015). Noradrenaline and dopamine neurons in the reward/effort trade-off: A direct electrophysiological comparison in behaving monkeys. Journal of Neuroscience, 35(20), 7866–7877.

Vogel, T. A., Savelson, Z. M., Otto, A. R., & Roy, M. (2020). Forced choices reveal a trade-off between cognitive effort and physical pain. Elife, 9, e59410.

Walton, M. E., & Bouret, S. (2019). What is the relationship between dopamine and effort? Trends in Neurosciences, 42(2), 79–91.

Watson, D., Clark, L. A., & Tellegen, A. (1988). Development and validation of brief measures of positive and negative affect: The PANAS scales. Journal of Personality Social Psychology, 54(6), 1063.

Westbrook, A., & Braver, T. S. (2015). Cognitive effort: A neuroeconomic approach. Cognitive, Affective, & Behavioral Neuroscience, 15(2), 395–415.

Westbrook, A., & Braver, T. S. (2016). Dopamine does double duty in motivating cognitive effort. Neuron, 89(4), 695–710.

Westbrook, A., Kester, D., & Braver, T. S. (2013). What is the subjective cost of cognitive effort? Load, trait, and aging effects revealed by economic preference. PloS One, 8(7), e68210.

Westbrook, A., van den Bosch, R., Määttä, J., Hofmans, L., Papadopetraki, D., Cools, R., & Frank, M. (2020). Dopamine promotes cognitive effort by biasing the benefits versus costs of cognitive work. Science, 367(6484), 1362–1366.

Zénon, A., Devesse, S., & Olivier, E. (2016). Dopamine manipulation affects response vigor independently of opportunity cost. Journal of Neuroscience, 36(37), 9516–9525.

