## supplemental_results for "Dopaminergic medication increases motivation to exert cognitive control by reducing subjective effort costs in Parkinson’s patients"

**Supplement**

In addition to participants choices in the DST, we also analyzed the time it took them to move the mouse cursor over the chosen color patch (Figure S1). Patients OFF dopamine were overall slower than healthy controls ($p$ = .002) and, marginally, than patients ON dopamine ($p$ = .066). This difference remained constant over the course of a block (disease state × trial number interaction: $p$ = .709; drug state × trial number interaction: $p$ = .183) and testing sessions (disease state × session interaction: $p$ = .952; drug state × session interaction: $p$ = .540). Participants with higher scores in the Symbol Digit Modalities Test (SDMT) responded faster in all groups (main effect SDMT: $p$ = .011; disease state × SDMT interaction: $p$ = .841; drug state × trial number interaction: $p$ = .426). For full results see Table S1.

| **Predictor** | ***b*** **(*SE*)** | ***p*** **- value** | |
|---|---|---|---|
| Intercept | -0.015 (0.030) | .623 | |
| Disease State | -0.161 (0.050) | .002 | ** |
| Drug state | -0.047 (0.025) | .066 | |
| Trial number | -0.001 (0.001) | .016 | * |
| Session | -0.039 (0.028) | .172 | |
| SDMT score | -0.064 (0.024) | .011 | * |
| Disease state × trial number | -0.001 (0.001) | .709 | |
| Drug state × trial number | -0.001 (0.001) | .183 | |
| Disease state × session | -0.002 (0.029) | .952 | |
| Drug state × session | 0.030 (0.049) | .540 | |
| Disease state × SDMT score | 0.011 (0.052) | .841 | |
| Drug state × SDMT score | -0.018 (0.022) | .426 | |

**Table S1.** Results of a Mixed-Effects Linear Regression predicting reaction times for color patch choices in the DST from disease state, drug state, trial number, session and SDMT score. SDMT = Symbol Digit Modalities Test; *** $p < .001$, ** $p < .01$, * $p < .05$

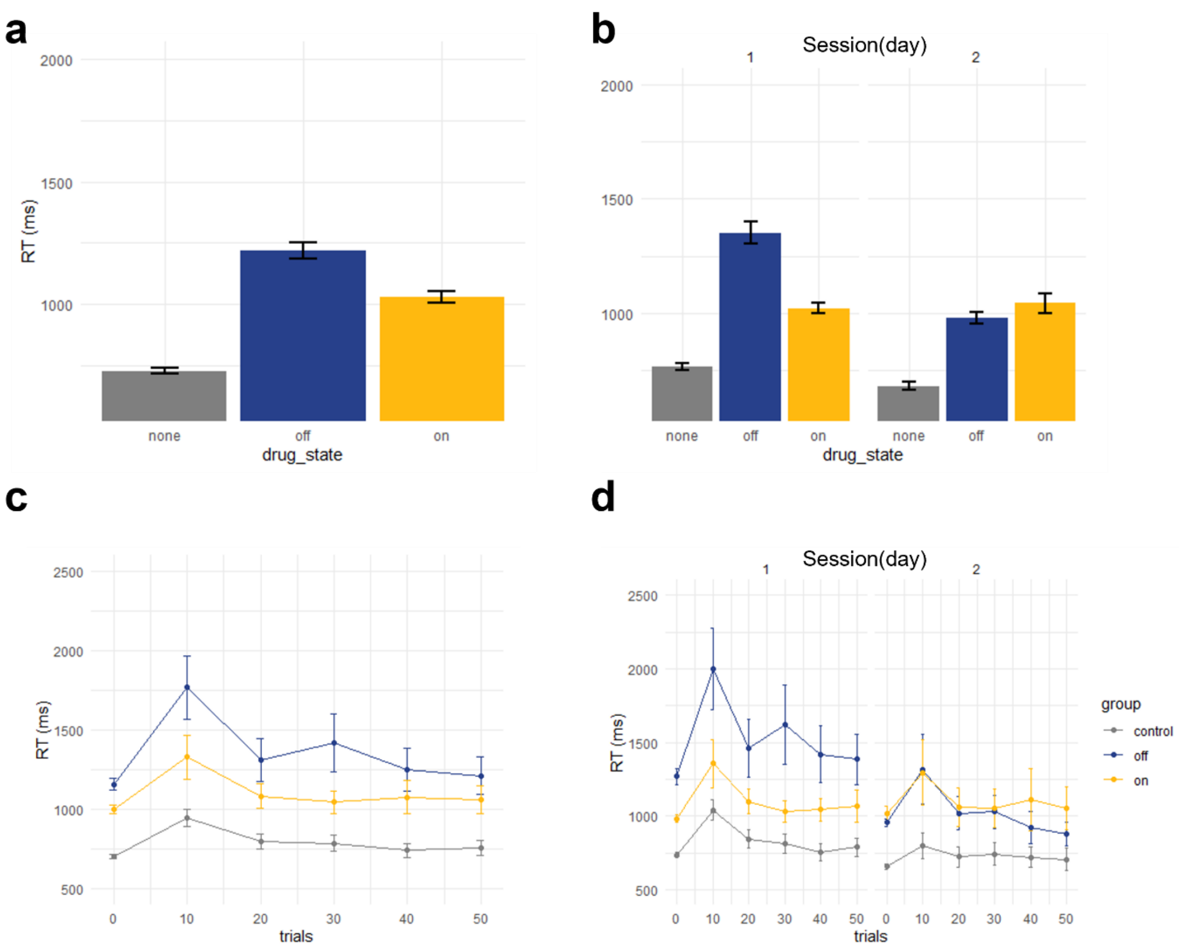


**Fig S1.** Choice reaction time in the free-choice blocks of the DST. Control participants were overall faster than PD patients, with patients OFF dopamine being slower than ON dopamine (a). The difference between medication state seemed to be weaker in session two (b). All participants became slightly faster in their choices across trials both overall (c) and in each session (d).

Given the prevalence of severe amotivation in PD and major depression (Husain & Roiser, 2018) as well as earlier reports suggesting a correlation between apathy ratings and reduced willingness to exert effort for reward (McGuigan et al., 2019), we also analyzed whether individual differences in the symptoms of apathy or depression, measured by the Apathy Evaluation Scale (AES) and the Geriatric Depression Scale (GDS), may predict choice behavior in the DST (Figure S2). However, we did not find a significant relation between questionnaire scores and effort avoidance in OFF patients (main effect AES: $p$ = .147; main effect GDS: $p$ = .076), ON patients (drug state × AES: $p$ = .921; drug state × GDS: $p$ = .310), or in healthy controls (disease state × AES: $p$ = .091; disease state × GDS: $p$ = .453). For full results see Table S2.

| | AES | | | GDS | | |
|---|---|---|---|---|---|---|
| **Predictor** | ***b* (*SE*)** | ***p* - value** | | ***b* (*SE*)** | ***p* - value** | |
| Intercept | 0.305 (0.114) | .007 | * | 0.285 (0.115) | .013 | * |
| Disease state | -0.112 (0.149) | .451 | | -0.074 (0.152) | .626 | |
| Drug state | -0.464 (0.157) | .003 | ** | -0.452 (0.159) | .005 | ** |
| Trial type | 0.001 (0.004) | .695 | | 0.002 (0.004) | .679 | |
| Session | 0.005 (0.110) | .963 | | -0.013 (0.111) | .908 | |
| Questionnaire score | -0.140 (0.096) | .147 | | 0.182 (0.102) | .076 | |
| Disease state × trial number | 0.001 (0.006) | .905 | | 0.001 (0.006) | .898 | |
| Drug state × trial number | 0.009 (0.003) | .001 | ** | 0.009 (0.003) | .001 | ** |
| Disease state × session | 0.020 (0.150) | .894 | | -0.020 (0.148) | .891 | |
| Drug state × session | -0.119 (0.142) | .400 | | -0.123 (0.146) | .397 | |
| Disease state × questionnaire score | 0.230 (0.136) | .091 | | -0.112 (0.149) | .453 | |
| Drug state × questionnaire score | 0.014 (0.137) | .921 | | -0.145 (0.143) | .310 | |

**Table S2.** Results of a Mixed-Effects Logistic Regression predicting proportion of choices according to instructions in the forced-choice blocks of the DST from disease state, drug state, trial number, session, and questionnaire scores. AES = Apathy Evaluation Scale, GDS = Geriatric Depression Scale; *** $p$ < .001, ** $p$ < .01, * $p$ < .05

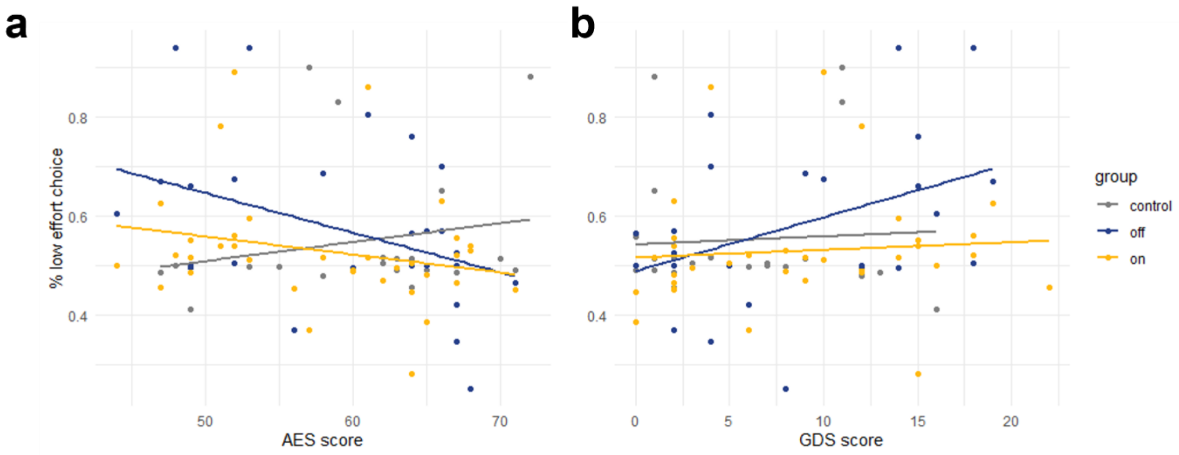


**Fig S2.** Relationship between questionnaire scores and effort avoidance. Individual differences in symptoms of apathy (a) and depression (b) do not predict proportions of low-demand choices. AES = Apathy Evaluation Scale. GDS = Geriatric Depression Scale.

**References**

Husain, M., & Roiser, J. P. (2018). Neuroscience of apathy and anhedonia: A transdiagnostic approach. *Nature Reviews Neuroscience*, *19*(8), 470.

McGuigan, S., Zhou, S.-H., Brosnan, M. B., Thyagarajan, D., Bellgrove, M. A., & Chong, T. T. (2019). Dopamine restores cognitive motivation in Parkinson’s disease. *Brain*, *142*(3), 719–732.
